# Comparison of logistic regression with regularized machine learning methods for the prediction of tuberculosis disease in people living with HIV: cross-sectional hospital-based study in Kisumu County, Kenya

**DOI:** 10.1101/2023.08.17.23294212

**Authors:** James Orwa, Patience Oduor, Douglas Okelloh, Dickson Gethi, Janet Agaya, Albert Okumu, Steve Wandiga

**Author notes:** **Corresponding author** James Orwa, Department of Population Health Sciences, Aga Khan University Kenya, P.O. Box 30270 – 00100, Nairobi, Kenya. **Email:** /.

## Abstract

**Background:** Tuberculosis (TB) is a major public health concern, particularly among people living with the Human immunodeficiency Virus (PLWH). Accurate prediction of TB disease in this population is crucial for early diagnosis and effective treatment. Logistic regression and regularized machine learning methods have been used to predict TB, but their comparative performance in HIV patients remains unclear. The study aims to compare the predictive performance of logistic regression with that of regularized machine learning methods for TB disease in HIV patients.

**Methods:** Retrospective analysis of data from HIV patients diagnosed with TB in three hospitals in Kisumu County (JOOTRH, Kisumu sub-county hospital, Lumumba health center) between [dates]. Logistic regression, Lasso, Ridge, Elastic net regression were used to develop predictive models for TB disease. Model performance was evaluated using accuracy, and area under the receiver operating characteristic curve (AUC-ROC).

**Results:** Of the 927 PLWH included in the study, 107 (12.6%) were diagnosed with TB. Being in WHO disease stage III/IV (aOR: 7.13; 95%CI: 3.86-13.33) and having a cough in the last 4 weeks (aOR: 2.34;95%CI: 1.43-3.89) were significant associated with the TB. Logistic regression achieved accuracy of 0.868, and AUC-ROC of 0.744. Elastic net regression also showed good predictive performance with accuracy, and AUC-ROC values of 0.874 and 0.762, respectively.

**Conclusions:** Our results suggest that logistic regression, Lasso, Ridge regression, and Elastic net can all be effective methods for predicting TB disease in HIV patients. These findings may have important implications for the development of accurate and reliable models for TB prediction in HIV patients.

## Introduction

Tuberculosis (TB) is a significant health burden in many parts of the world, particularly in people living with Human Immunodeficiency Virus (PLWHIV), who are at increased risk of developing the disease. In 2021, about 187 000 people died of HIV-associated TB related conditions, with the 76 percent of notified TB patients tested for HIV test and knew their result. The percentage of notified TB cases was an increase from 73 percent reported in 2020(1). To achieve the 2030 global tuberculosis mortality reduction of 90% and 80% incidence (SDG targets goal 3.3), early and accurate diagnosis of TB disease of all types including drug sensitive, and drug resistant TB is important(2).

Africa bears high burden of TB and HIV co-infection as 10.6 million people infected with TB in 2021 globally, 23% were in Africa and 6.7% of them were coinfected with HIV(3). Kenya is one of the 30 countries in SSA with the high TB burden in the world and the fifth in Africa(3). The estimated HIV prevalence as reported by Kenya Indicator Aids Survey of 2012 was 7.1% with the highest burden in Nyanza province of 15.1%(4) . The Kenya Population-based Impact Assessment (KENPHIA) survey reported a national HIV prevalence of 4.9% which was lower the prevalence of 2012 survey(5). Kisumu county is third in Kenya among counties with the highest burden of HIV after Siaya and Homabay counties(6). HIV is driving the TB epidemic in areas where there is high HIV prevalence ((7). It is due to this TB/HIV synergy that World Health Organization(WHO) recommendation that every HIV patient should be screened for TB clinical symptoms of cough, fever, night sweats and weight loss and vice versa (8).

The highest priority for TB control is the identification and treatment of infectious cases identified by at least sputum smear, culture tests or clinical symptoms among the population at risk. One of the ways to identify infected cases is through intensive case finding especially in countries with high TB burdens. Intensive case finding primarily involves detecting TB among symptomatic patients who present to health facilities for HIV care and treatment services. The treatment of TB among people living with HIV (PLWH) has a high pill burden, drug toxicity, drug-drug interactions and, tuberculosis associated immune reconstitution inflammatory syndrome and these remains a challenge to the treatment especially in resource-limited settings(9).

Factors found to be influencing TB among PLWH had been explored in several studies and includes age of the respondents, baseline CD4 count cells (10), WHO disease stage 3 and 4 (2, 7) and previous history of TB disease(11). The analysis of these factors were based on traditional logistic regression models and has not benefited from newer techniques such as machine learning technique(9, 12, 13). Early and accurate prediction of TB in PLWH using machine learning techniques is crucial for effective treatment and control of the disease. Logistic regression (LR) is a popular statistical method used for binary classification tasks such as disease prediction. However, LR assumes linearity and independence of predictors, which may not be realistic in real-world data. Regularized machine learning methods such as Lasso, Ridge, and Elastic net regression have been proposed as alternatives to LR to improve prediction accuracy and handle high-dimensional data. In this study, we compare the performance of LR with Lasso, Ridge, and Elastic net regression for the prediction of TB in HIV patients using a cross-sectional hospital-based dataset. Our results provide insights into the suitability of these methods for predicting TB in HIV patients, which can aid in the development of effective screening programs and treatment strategies.

## Methodology

### Study design and setting

This was an observational hospital-based prospective study among PLWH accessing care and treatment service in three highly enrolling HIV care and treatment centres in Kisumu, Kenya which were Jaramogi Oginga Odinga Teaching and Referral Hospital, Kisumu County referral hospital and Lumumba Health Centre between 9^th^ January 2014 to 15^th^ August 2017. Kisumu County is a county located in western Kenya, with its capital city Kisumu. It covers an area of approximately 2,085 square kilometres and has a population of over 1.2 million people. It is home to Lake Victoria, the largest freshwater lake in Africa and the source of the Nile River. The county is known for its rich cultural heritage, with the Luo people being the largest ethnic group. The main economic activities include fishing, agriculture, and trade. Kisumu is also a hub for transportation and communication in the region, with a modern airport, port, and railway station.

Kisumu County in Kenya has a high burden of HIV and Tuberculosis (TB) infections. According to the Kenya AIDS Response Progress Report 2021, the county had an HIV prevalence rate of 13.9%, which is higher than the national average. The report also indicated that approximately 62% of people living with HIV in Kisumu County are on antiretroviral therapy (ART) to manage the infection(14).

Regarding TB, Kisumu County is ranked among the high TB burden counties in Kenya. According to the National TB prevalence survey of 2016, Kisumu had a TB prevalence rate of 390 per 100,000 people, which is higher than the national average(15). The County Government of Kisumu, together with development partners and stakeholders, is implementing several strategies to control and manage the spread of both HIV and TB, including scaling up testing, treatment, and prevention services. The selection of the sites were informed by a collaborative meeting between Kenya Medical Research Institute (KEMRI)-Center for Global Health Research (CGHR)-TB branch and Ministry of Health. Participants who were on TB treatment at the time of the initial presentation to the clinic, being treated for TB diseases or infection within the one year preceding their initial presentation or without HIV positive documented result were excluded from the study.

### Variables

The primary outcome was TB diagnosed measured on a binary scale (positive, negative) based on any of the tests. A positive TB case was defined as active TB bacteriologically confirmed by smear microscopy, culture or Xpert tests. Demographic and clinical variables used as predictors included: Age in years, gender, smoking status, use of illegal drugs, alcohol consumption, ART initiation, baseline CD4 counts, WHO disease staging, baseline symptoms in the last four weeks (cough, fever, night sweats, and weight loss).

### Data collection tools and procedure

Eligible patients were informed about the study during their clinic days and invited to participate. Data on demographic were collected during the enrollment visit. Enrolled participants were clinically evaluated three times during the study (enrollment, 2-3 days later, third follow-up visit). The second visit was to determine and report the results of latent tuberculosis infection testing, whereas the third visit was for repeat clinical evaluation of the symptoms. The TB disease symptoms were screened using the WHO-recommended clinical screening tool which consists of questions on current cough, fever, night sweats, and weight loss at any time of the scheduled study visit.

### Specimen collection procedure

Regardless of the symptoms, each patient was asked to produce at least one sputum specimen at the time of initial visit (a spot specimen). This was tested using smear microscopy to diagnose TB. Participants whose initial smear microscopy were negative were requested to bring at least one additional specimen for testing, either a morning specimen (if they remember to bring it with them), or a second spot specimen (if they forgot to produce and bring the morning specimen) during the second visit. During the follow-up visit, two additional sputa were collected, one of which was the morning specimen or two spot produced at the time of the visit within one hour interval.

Initial samples collected were stored in a refrigerator or cooler box until they were transported to the central mycobacteriology testing TB laboratory at KEMRI CGHR in Kisian for processing and culture. Culture was performed in liquid media, using the Bactec Mycobacterial Growth Indicator Tube (MGIT) 960 system. Sputum specimen collected at later visits were sent directly to the central laboratory for smear microscopy and culture and testing with the GeneXpert MTB/RIF assay. The third visit specimen was tested by culture and Xpert MTB/RIF test using Cepheid’s GeneXpert platform. The Xpert MTB/RIF assay was used as an add-on diagnostic to detect rifampin-resistant strains.

Sputum culture was used as the gold standard for TB diagnosis. Any patients whose sputum smear shows acid fast bacilli (AFB) on microscopy, or whose sputum culture grows *M.TB*, were notified immediately to return to clinic as soon as possible for a follow- up clinical evaluation and to initiate TB treatment therapy as per the Kenyan Government Ministry of Health guidelines(16). On the other hand, all patients with negative culture results were considered for isoniazid preventive therapy as part of their routine HIV care.

### Data processing and analysis

Prior to model development, data were explored descriptive using frequency and percentage for categorical variables and median (interquartile range (IQR)). All variables were considered clinically important in explaining HIV infection and were included in the multivariable logistic regression modelling to assess independence and strength of association in terms of odds ratios.

We trained our dataset on a logistic, ridge, LASSO, and elastic net regressions to find which classifier maximized the accuracy of our TB disease prediction and to ensure they were robust across various feature selection methods. Due to the size of the dataset, there was no split of the data and all dataset were used as training dataset. Ten-fold cross validation with five repeats was used to develop the prediction models and to avoid model overfit. The tuning parameter grids used for ridge and lasso was set as 10^seq(2, -3, by = -.1) and elastic net set with a tuning length of ten. Age in years and CD4 results were normalized and missing CD4 results were imputed with the median values. The area under curve (AUC) and accuracy were used to compare the models generated by different methods. The relative importance of predictors was plotted using variable importance plot. The aim of the predictive modelling was not to test whether some variables affect another or to study causal relationships, instead the goal is to make good predictions for the TB disease. Each classifier is described below. The models were implemented in RStudio with the *caret* and *glmnet R* package.

### Logistic regression

Logistic regression (LR) is used principally for predicting binary or multi-class dependent variables given a set of predictors. This algorithm’s response variable is binary, and it builds a model to predict the odds of its occurrence. The limiting assumptions of normality and independence of this method have contributed to an increase in the application and popularity of machine learning techniques to real-world prediction problems. The logistic function is defined as

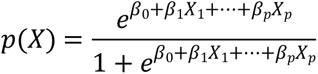

Where *β*_*i*_ parameters represent the parameter coefficients, *X*_*i*_ are the explanatory variables and p(X) is the probability of having TB disease.

### Ridge regression

This is a method of shrinkage that constrains the effect of irrelevant estimates to shrink to zero. The objective function is like logistic regression, however there is a penalty term added that controls the estimated coefficients by adding 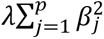 to the objective function. The *λ* is the tuning parameter. The ridge for the logistic regression minimizes the function(17).

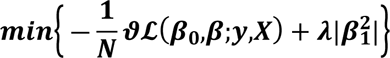

Where N=sample size

*β*_0_ = intercept

*β*=vector of logistic regression coefficients

Y=vector (length N) of outcomes

X=matrix of independent variables

*λ*=penalty parameter

### LASSO regression

LASSO is a means of feature selection to shrink the coefficients of “less predictive” covariates to zero. The least absolute shrinkage and selection operator (lasso) penalty is an alternative to the ridge that requires a small modification in the penalty. The penalty is defined as 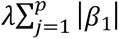. whereas the ridge penalty pushes variables to approximately but not equal to zero, the lasso penalty will push coefficients all the way to zero. Switching to the lasso penalty not only improves the model but it also conducts automated feature selection. The lasso penalty minimizes the following negative log-likelihood function(17).

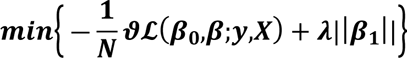

Where

N=sample size

*β*_0_ = intercept

*β*=vector of logistic regression coefficients

Y=vector (length N) of outcomes

X=matrix of independent variables

*λ*=penalty parameter

### Elastic net regression

This is a generalization of the ridge and lasso penalties that combines the two penalties. The advantage of elastic net penalty is that it enables effective regularization via the ridge penalty with the feature selection characteristics of the lasso penalty. Both methods reduce the variance hence improves the fit of the model. The elastic net estimator for logistic regression minimizes(17).

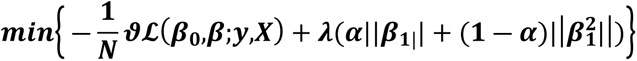

Where N=sample size

*β*_0_ = intercept

*β*=vector of logistic regression coefficients

Y=vector (length N) of outcomes

X=matrix of independent variables

*λ*=penalty parameter

### Ethical consideration

The study was approved by Kenya Medical Research Institute Scientific and Ethical Review Unit with approval reference number SSC#2670. Other ethical approvals were obtained from the respective hospitals and from the County Ministry of Health after being briefed about the purpose and benefits of the study. All patients provided written signed consent before the start of the data collection. Patients who were under the age of 18 were asked to sign an assent form, and their legal guardian signed a consent form on their behalf. The study was conducted in accordance with the ethical standards of the KEMRI institutional scientific research committee.

## Results

### Baseline characteristics

A total of 927 participants were screened, out of whom 893 were eligible, 849 participants consented to participate in the study and were enrolled. The median age was 32 (IQR: 26- 39) years. Majority of participants were females (60.8%, n=516), 25 (2.9%) were smokers, 10 (1.2%) were users of illegal drugs, and 53 (6.2%) were consuming alcohol. Slightly half (50.8%; n=431) of the participants were on ART, the median CD4 cell counts were 308 (IQR: 155.5-507.0), and 730 (86.0%) had been started on cotrimoxazole preventive therapy. The number in each WHO disease stage decreased as the stage advances with only 79 (9.3%) of the participants in WHO disease stage IV. TB disease was diagnosed in 107 (12.6%; 95%CI:10.4%-14.8%) of the patients (Table 1).

**Table 1:** Baseline demographic and clinical characteristics of the participants, N=849.

| Variable | N = 849<sup>1</sup> |
| --- | --- |
| Age in years, Median (IQR) | 32.0 (26.0 – 39.0) |
| Gender, n (%) |  |
| Males | 333 (39.2) |
| Females | 516 (60.8) |
| Smoking, n (%) | 25 (2.9) |
| Use of any illegal Drugs, n (%) | 10 (1.2) |
| Consume alcohol, n (%) | 53 (6.2) |
| Started on ART, n (%) | 431 (50.8) |
| Baseline CD4 count, Median (IQR) | 308.0 (155.5 – 507.0) |
| Started on CPT, n (%) | 730 (86.0) |
| WHO stage, n (%) |  |
| WHO stage 1 | 430 (50.6) |
| WHO stage 2 | 340 (40.0) |
| WHO stage 3/4 | 79 (9.3) |
| TB status |  |
| Negative | 742 (87.4) |
| Positive | 107 (12.6) |

### Presenting symptoms at baseline and follow-up

There was a reduction in the number of symptoms during the follow-up period compared to those presented at baseline. Weight loss was reported by most of the participant baseline, whereas cough in the last four weeks was the most common symptoms at the time of follow-up. There were 141 (15.6%) who reported all the symptoms at baseline, this reduced to only 30 (3.6%) of the patients at follow-up (Table 2).

**Table 2:** Presenting symptoms at enrollment and follow-up visits.

| Symptoms in the last 4 weeks | Visit type |  |
| --- | --- | --- |
|  | Enrollment, N = 849 | Follow-up, N = 836 |
| Cough, n (%) | 412 (48.5) | 210 (25.1) |
| Fevers, n (%) | 408 (48.1) | 178 (21.3) |
| Night sweats, n (%) | 323 (38.0) | 148 (17.7) |
| Weight loss, n (%) | 454 (53.5) | 176 (21.1) |
| All symptoms, n (%) | 141 (16.6) | 30 (3.6) |

### Determinants of TB disease

Only WHO disease stage III/IV and cough in the last four weeks during baseline were significantly associated with the TB disease in PLWH in the multivariable analysis. The odds of getting TB disease among those who had advanced HIV disease stage was seven time higher than the participants in WHO disease stage I (aOR=7.13; 95%CI:3.86-13.33). Individuals who reported a cough in the past four weeks were found to have more than double the odds of developing tuberculosis compared to those without a recent cough (aOR:2.34; 95%CI:1.43-3.89) (Table 3).

**Table 3:** Factors Associated with the Likelihood of TB Diagnosis in PLWH: Results from a Logistic Regression Analysis.

| Variables | TB diagnosis |  | Univariate |  | Multivariable |  |
| --- | --- | --- | --- | --- | --- | --- |
|  | Negative | Positive | OR (95%CI) | P-value | aOR (95%CI) | P-value |
| Age, median (IQR) | 31.0 (26.0-39.0) | 35.0 (29.0-39.0) | 1.01 (0.99-1.03) | 0.233 | 1.00 (0.98-1.03) | 0.697 |
| Gender |  |  |  |  |  |  |
| Males | 280 (84.1) | 53 (15.9) |  |  |  |  |
| Females | 462 (89.5) | 54 (10.5) | 0.62 (0.41-0.93) | 0.02 | 0.76 (0.47-1.21) | 0.238 |
| Currently smoking |  |  |  |  |  |  |
| No | 720 (87.4) | 104 (12.6) |  |  |  |  |
| Yes | 22 (88.0) | 3 (12.0) | 0.94 (0.22-2.79) | 0.972 | 0.81 (0.17-2.85) | 0.763 |
| Use illegal drugs |  |  |  |  |  |  |
| No | 733 (87.4) | 106 (12.6) |  |  |  |  |
| Yes | 9 (90.0) | 1 (10.0) | 0.77 (0.04-4.15) | 0.804 | 0.75 (0.04-5.39) | 0.802 |
| Consume Alcohol |  |  |  |  |  |  |
| No | 697 (87.6) | 99 (12.4) |  |  |  |  |
| Yes | 45 (84.9) | 8 (15.1) | 1.25 (0.53-2.60) | 0.573 | 1.50 (0.59-3.38) | 0.359 |
| ART |  |  |  |  |  |  |
| No | 359 (85.9) | 59 (14.1) |  |  |  |  |
| Yes | 383 (88.9) | 48 (11.1) | 0.76 (0.51-1.14) | 0.192 | 0.80 (0.49-1.31) | 0.374 |
| CPT |  |  |  |  |  |  |
| No | 98 (82.4) | 21 (17.6) |  |  |  |  |
| Yes | 644 (88.2) | 86 (11.8) | 0.62 (0.38-1.07) | 0.076 | 0.59 (0.32-1.10) | 0.09 |
| WHO stage |  |  |  |  |  |  |
| I | 394 (91.6) | 36 (8.4) |  |  |  |  |
| II | 305 (89.7) | 35 (10.3) | 1.26 (0.77-2.05) | 0.361 | 1.14 (0.69-1.90) | 0.604 |
| III/IV | 43 (54.4) | 36 (45.6) | 9.16 (5.25-16.12) | < 0.001 | 7.13 (3.86-13.33) | < 0.001 |
| Cough in the last 4 weeks |  |  |  |  |  |  |
| No | 409 (93.6) | 28 (6.4) |  |  |  |  |
| Yes | 333 (80.8) | 79 (19.2) | 3.47 (2.23-5.54) | < 0.001 | 2.34 (1.43-3.89) | 0.001 |
| Fever last 4 weeks |  |  |  |  |  |  |
| No | 408 (92.5) | 33 (7.5) |  |  |  |  |
| Yes | 334 (81.9) | 74 (18.1) | 2.74 (1.79-4.28) | < 0.001 | 1.54 (0.91-2.62) | 0.109 |
| Night sweets |  |  |  |  |  |  |
| No | 473 (89.9) | 53 (10.1) |  |  |  |  |
| Yes | 269 (83.3) | 54 (16.7) | 1.79 (1.19-2.70) | 0.005 | 0.71 (0.43-1.18) | 0.191 |
| Weight loss |  |  |  |  |  |  |
| No | 369 (93.4) | 26 (6.6) |  |  |  |  |
| Yes | 373 (82.2) | 81 (17.8) | 3.08 (1.96-4.99) | < 0.001 | 1.83 (1.09-3.14) | 0.24 |

The first and second vertical dashed lines represent the λ value with the largest AUC and the largest λ value within one standard error of the largest AUC that gives the best ROC-AUC. In both cases the AUC increase slightly with an increase in the *λ* and starts to drop when the value of AUC is reaches its peak. The maximum value of AUC is reached when the value for *λ* that produces an optimal trade-off between bias and variance is reached. Similarly, the number of parameters shrinks with the increasing *λ* to avoid overfitting of the two models.

**Figure 1:**
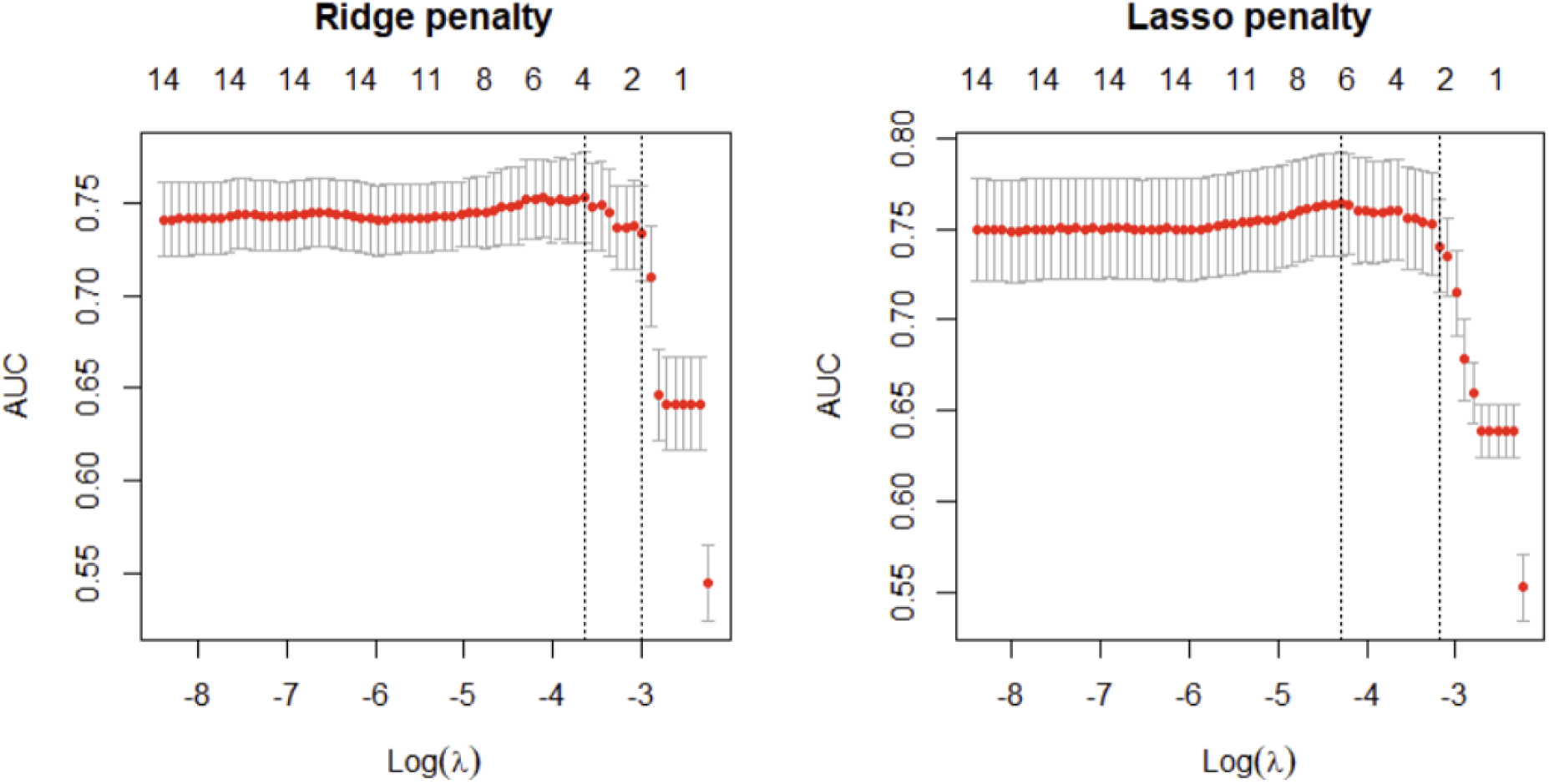
Ridge and LASSO models showing AUC values with the corresponding *λ* values that gives the best AUC values.

### Comparing the performance of the algorithms

All the regularization models achieved statistically improvements in AUC compared to the standard logistic regression model. The AUC of ridge regression with minimum *λ* =0.02616 was 0.753% and for LASSO with minimum *λ*=0.01264 was 0.764%. Logistic regression and elastic net regression had accuracy of 0.868% and 0.874% respectively. Elastic net model performed well in predicting TB disease out of the other models trained.

| Models | Accuracy | AUC |
| --- | --- | --- |
| LR | 0.868 | 0.744 |
| Ridge | 0.868 | 0.753 |
| LASSO | 0.869 | 0.764 |
| Elastic net | 0.874 | 0.762 |

The variable importance graph for the best model is shown in figure **XX.** Night sweets in the last four weeks and use of alcohol were not important predictors and were excluded in the model. The most important predictor in the model was WHO disease stage III/IV, followed by cough in the last four weeks. Participants age and use of illegal drugs were least important predictors in the predictive model.

**Figure 2:**
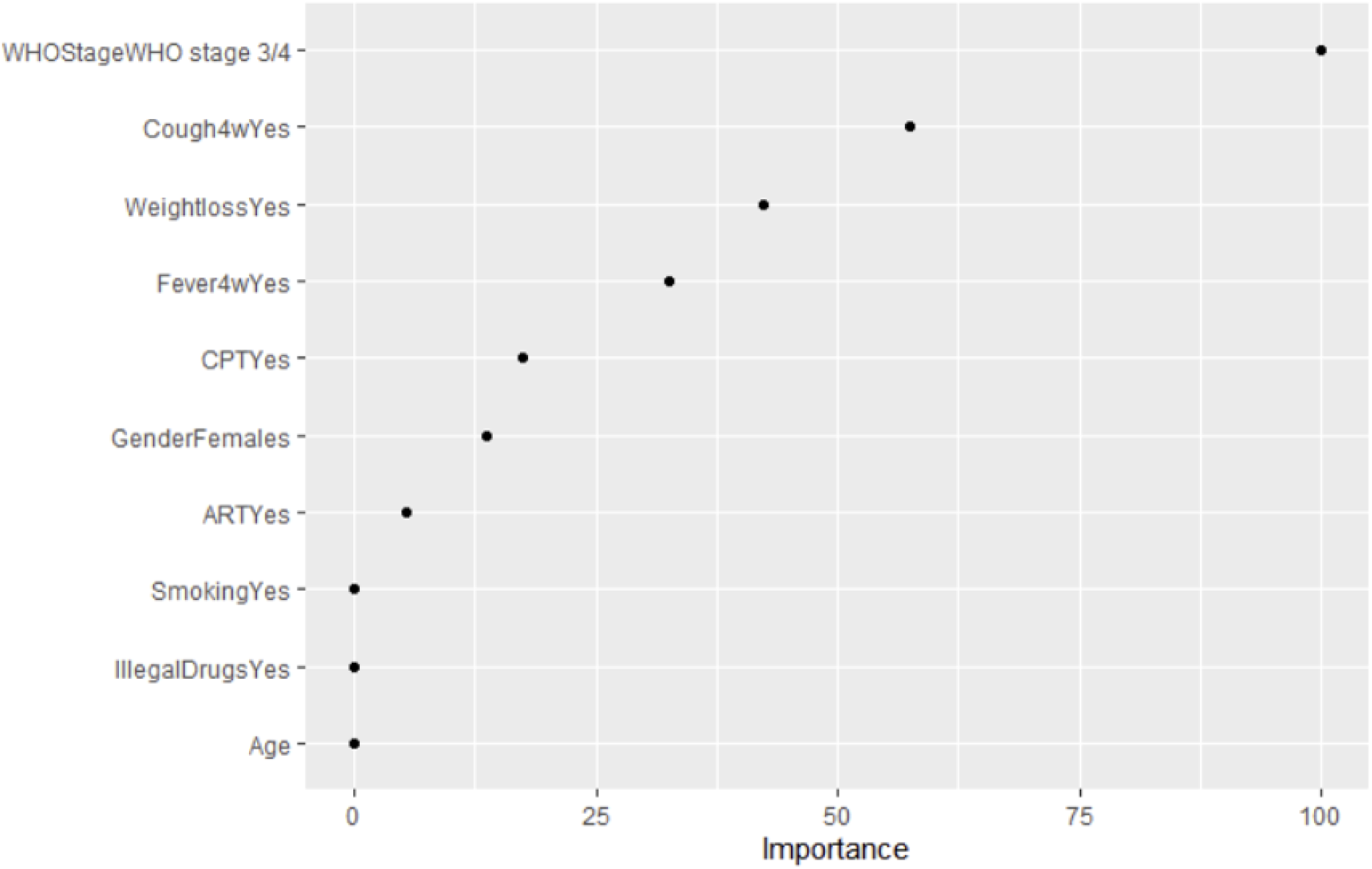
Variable arranged by order of importance in predicting TB disease in PLWH.

## Discussion

TB remains to be a major opportunistic infection and contributes to high mortality among PLWH. The diagnosis of TB in PLWH is a major public health problem. The WHO symptom screening algorithm for the detection of pulmonary TB among PLWH needed laboratory confirmation particularly in settings with limited resources to implement the guidelines. The current algorithm recommends that PLWH are screened by assessing symptoms and by using tests, examinations or other procedures that can be applied rapidly and if found positive to be referred for laboratory tests. In this analysis, we set to find out which predictors explains the disease and to compare the predictive performance of different models.

In this study we found that patients in WHO stage III/IV and those with symptoms of cough were more likely to test positive with the TB disease. Use of cough symptom as a predictor of TB had shown high sensitivity among PLWH in Ethiopia compared to the other TB symptoms (18). Coughing was also associated with high odds of being infected with TB in a large national survey in Kenya(19).However, this was not the case in the general population as found in a study in south Africa(20). Our findings on the advanced WHO staging were similar to the findings among newly diagnosed HIV individuals in western Kenya that found three-fold likelihood of TB disease among this population of newly HIV diagnosed individuals(21).

We compared the performance of logistic regression with regularized machine learning methods for predicting the occurrence of TB in PLWH. Our results show that the regularized machine learning methods outperformed the logistic regression model in terms of both accuracy and area under the receiver operating characteristic curve (AUC-ROC). The best performing method was the elastic net regression, which achieved an AUC of 0.76, compared to 0.74 for the logistic regression model.

Our findings suggest that regularized machine learning methods may be more effective for predicting TB in HIV patients than traditional logistic regression models. This is likely due to the ability of regularized machine learning methods to handle high-dimensional data and avoid overfitting, which can be a significant problem in logistic regression models. Regularized machine learning methods are also more flexible in terms of the types of data they can handle and can incorporate complex relationships between variables.

Our study has several limitations, including the cross-sectional design and the limited sample size. We also did not include external validation of our models, which could limit the generalizability of our findings to other populations. Future studies should consider longitudinal designs and larger sample sizes to further explore the performance of these models in predicting TB in HIV patients.

In conclusion, our study provides evidence that regularized machine learning methods, particularly the elastic net regression model, may be more effective than logistic regression in predicting the occurrence of TB in HIV patients. These findings have important implications for clinical decision-making and the development of more accurate and effective diagnostic tools for TB in PLWH especially among individuals with advanced WHO disease stage and coughing.

## Data Availability

Data cannot be shared publicly because of ethical issues. Data are available from the KEMRI Institutional Data Access / Ethics Committee (contact via) for researchers who meet the criteria for access to confidential data.

## Acknowledgement

We would like to thank the study participants, the study team, and collaborating partners from the ministry of health, and the health facilities in-charges of the participating health facilities

